# Tick-Induced Mammalian Meat Allergy in Australia: National Prevalence and Geographic Distribution from Laboratory Surveillance, 2014-2024

**DOI:** 10.64898/2026.07.13.26358004

**Authors:** Emily Smith, Paul Campbell, Carl Kennedy, Karl Baumgart, Lucinda Wallman, Stephen C. Barker, Sheryl van Nunen, Andrew A. Walker, Alexander W. Gofton

## Abstract

**Objectives:** To characterise the national epidemiology of tick-induced mammalian meat allergy (MMA) in Australia, including temporal trends in α-Gal specific IgE testing and case detection, geographic distribution, demographic risk factors, and longitudinal antibody dynamics.

**Design:** Retrospective analysis of α-Gal sIgE ImmunoCAP™ test results from 1 January 2014 to 31 December 2024.

**Setting, Participants:** Deidentified laboratory records from 14,075 individuals tested across Australia, with residential postcodes mapped to Statistical Areas Level 3 (SA3). A subset of 1,515 individuals with repeat testing contributed to longitudinal analyses.

**Main outcome measures:** Test volumes, suspected MMA case counts, positivity rates, spatial clustering metrics, demographic risk ratios, and longitudinal α-Gal sIgE trajectories.

**Results:** Overall, 35.7% (5,025) of individuals tested positive. Testing volume increased 331% over the study period, with case detection accelerating sharply from 2020. Decomposition analysis attributed 59–81% of case growth to expanded testing, with the remainder unexplained by surveillance expansion alone. Cases were concentrated along the eastern seaboard within the range of *Ixodes holocyclus*, with extreme spatial clustering: three SA3 regions accounted for over one-quarter of national cases. Females comprised the majority of positive cases, and MMA risk increased with age, peaking at 45–74 years. Among 1,515 individuals with serial testing, α-Gal sIgE levels antibodies declined predictably in 93% of people.

**Conclusions:** This first national assessment of MMA in Australia reveals a substantial, geographically concentrated, and growing burden. The extreme spatial concentration of cases suggests that targeted public health interventions could efficiently address a large proportion of national disease burden. While testing expansion is the dominant driver of rising case numbers, it does not fully account for observed trends, and prospective studies are needed to disentangle surveillance effects from genuine disease emergence. Declining antibody levels support the utility of serial testing for clinical monitoring, though persistent sensitisation underscores the need for sustained risk mitigation and continued surveillance.

**Plain Language Summary Box:** *The known:* In Australia, bites from the paralysis tick can trigger mammalian meat allergy (MMA), causing delayed allergic reactions to red meat. The national scale of this condition was previously unknown.

*The new:* We analysed MMA laboratory data over 11 years. Cases have risen sharply since 2020, faster than increased testing alone can explain. Most occur along the east coast, with just nine regions accounting for half of all cases. Middle-aged and older adults, particularly women, are most affected.

*The implications:* The geographic concentration of cases in few regions suggests targeted public health interventions could efficiently reduce national disease burden.

## 1. Introduction

Mammalian meat allergy (MMA), also known as alpha-Gal syndrome, is an IgE-mediated allergic condition characterised by delayed-onset hypersensitivity reactions to galactose-α-1,3-galactose (α-Gal), an oligosaccharide abundant in non-primate mammalian tissues. Unlike typical food allergies, MMA is distinguished by a characteristic 3–8-hour delay between mammalian meat consumption and symptom onset, attributed to the time required for digestion of α-Gal–containing glycolipids [1]. The clinical spectrum of MMA ranges from mild urticaria and gastrointestinal symptoms to life-threatening anaphylaxis, with severe reactions reported in up to 60% of affected individuals [2]. Beyond mammalian meat, reactions may occur following exposure to mammalian-derived products including dairy, gelatine, and pharmaceuticals or medical devices bearing α-Gal such as cetuximab and porcine heart valves [3]. Australia’s first confirmed MMA fatality was confirmed posthumously in February 2026, following the first reported fatal case in the United States in 2025 [4, 5]. MMA presents significant diagnostic challenges due to its delayed reaction, variable clinical manifestations, and lack of clinician and public awareness of the condition [6, 7].

MMA is induced by tick (Acari: Ixodidae) bites, with cases reported in at least 50 countries [8]. MMA has been attributed to various tick species worldwide, with the eastern paralysis tick (*Ixodes holocyclus*) the established causative species in Australia [9, 10]. *Ixodes holocyclus* is distributed along Australia’s eastern seaboard, co-occurring with approximately 50% of Australia’s population, including major population centres in south-east Queensland (QLD) and New South Wales (NSW) [11, 12]. Within its range, *I. holocyclus* accounts for almost all reported human tick bites due to its generalist host preference and prevalence in peri-urban residential settings and outdoor recreation areas [12, 13].

Our understanding of MMA has advanced substantially in recent years, though significant knowledge gaps remain, particularly in Australia, where comprehensive epidemiological data are lacking. Previous estimates of MMA prevalence in Australia were limited to a single region (approximately 113 cases per 100,000 in the Sydney Basin), derived from clinical cohorts rather than systematic surveillance [8]. Consequently, the true burden of MMA in Australia remains poorly characterised, hampering clinical recognition and leaving clinicians, public health authorities, and policymakers without the evidence base needed to respond to this emerging health threat.

This study addresses these gaps by analysing α-Gal-specific IgE (sIgE) testing data from Australian pathology providers over an 11-year period (2014–2024, inclusive). We sought to characterise the burden and geographic distribution of MMA nationally, identify demographic risk factors, examine temporal trends in testing and case detection, and describe longitudinal changes in α-Gal sIgE levels among patients with repeated testing. These data provide the first comprehensive epidemiological assessment of MMA in Australia and establish baseline metrics for ongoing surveillance of this emerging allergic condition.

## 2. Methods

### 2.1 Data collection and processing

Deidentified data were obtained from all α-Gal sIgE ImmunoCAP™ (Phadia AB, Uppsala, Sweden) tests submitted to QML Pathology, Sullivan & Nicolaides Pathology, Douglass Hanly Moir Pathology, and Laverty Pathology between January 1, 2014, and December 31, 2024. These laboratories provide services across all Australian states and territories and conduct the majority of α-Gal sIgE testing in Australia. Although the exact proportion of total national testing these providers represent is unknown, the authors were unable to find other public or private pathology services that offered this test. Each record contained a unique deidentified patient and test identifier, year of birth, sex, residential postcode, date of test, and the test result in kilounits of α-Gal sIgE (kU/L). Clinical records, travel histories, and any other tests results were not obtained. Data with invalid postcodes or those from outside Australia or missing essential information were excluded (*n*=5).

Patients with at least one positive result (α-Gal sIgE ≥ 0.1 kU/L) were classified as suspected MMA cases. In the absence of clinical data, this classification acknowledges that a positive result provides supportive but not confirmatory evidence of MMA and assumes that testing was requested based on clinical suspicion of MMA. However, we acknowledge that test indications may have varied and could include investigation of idiopathic anaphylaxis or tick-bite reactions. For patients receiving multiple positive tests demographic and location data were taken from their first positive test.

Residential postcodes were aggregated to Australian Bureau of Statistics (ABS) Statistical Areas Level 3 (SA3) regions (Australian Statistical Geography Standard Edition 3) using spatial intersection analysis, assigning each postcode to the SA3 with the greatest geographic overlap using the *sf* package in R v4.4 [14, 15]. SA3 regions represent functional population areas of 30,000–130,000 people, providing a meaningful unit for regional comparison. Suspected MMA cases were expressed as cases per million population per year (PPY) for each SA3 region, using annual ABS regional population estimates as denominators.

### 2.2 Statistical analysis

Temporal trends in testing volume and case numbers were characterised using segmented regression to identify breakpoints in growth trajectories. Demographic risk ratios were calculated by age, sex, and age-sex combinations. Geographic analyses quantified spatial heterogeneity in MMA burden using the Gini coefficient and spatial autocorrelation using Moran’s I. To quantify the extent to which increased case detections reflected expanded testing versus genuine changes in sensitisation prevalence, we decomposed case growth into volume, positivity rate, and interaction components using the pooled 2018–2020 period as the reference baseline. This decomposition was extended to separate geographic expansion (new SA3 regions tested) from testing intensification within existing regions. Longitudinal antibody dynamics were modelled using linear mixed-effects models to account for repeated measures within patients. All analyses used R v4.4.1. Statistical tests are specified in Results with corresponding p-values; two-sided p < 0.01 was considered significant unless otherwise noted. The full analytic workflow, including all code used in this study, is available at https://github.com/alexandergofton/Tick-Induced_MMA_in_Australia.

### 2.3 Ethics

This study was approved by the CSIRO Health and Medical Research Ethics Committee (approval #2025_003_LR). As this study used retrospectively collected, deidentified laboratory data with no patient contact, individual consent was not required.

## 3. Results

### 3.1 Patient population and demographic risk factors

From 1 January 2014 to 31 December 2024, 16,562 α-Gal sIgE ImmunoCAP™ tests were performed on 14,075 people, with just 10.8% (1,515) of people receiving multiple tests (range 2-15). Over the study period, 35.7% (5,025) of people tested were positive for α-Gal sIgE (≥ 0.01kU/L) and were classified as suspected MMA cases. Females predominated among those tested (64.8%) and positive cases (58.6%) **(Table 1**; **Figure 1)**. Testing volume peaked in the middle age (median age 48-49 years; **Table 1**; **Figure 1A**), with suspected MMA cases being moderately older than those testing negative (Wilcoxon: p < 0.01; Kolmogorov-Smirnov: p < 0.01; Cohen’s d = 0.33), with peak positivity occurring at ages 55-74 for males and 45-74 for females **(Table 1**; **Figure 1B)**. Despite lower testing rates, males had overall higher positivity rates than females (42.0% vs 32.3%) **(Tables 1, S1)**.

**Table 1.**
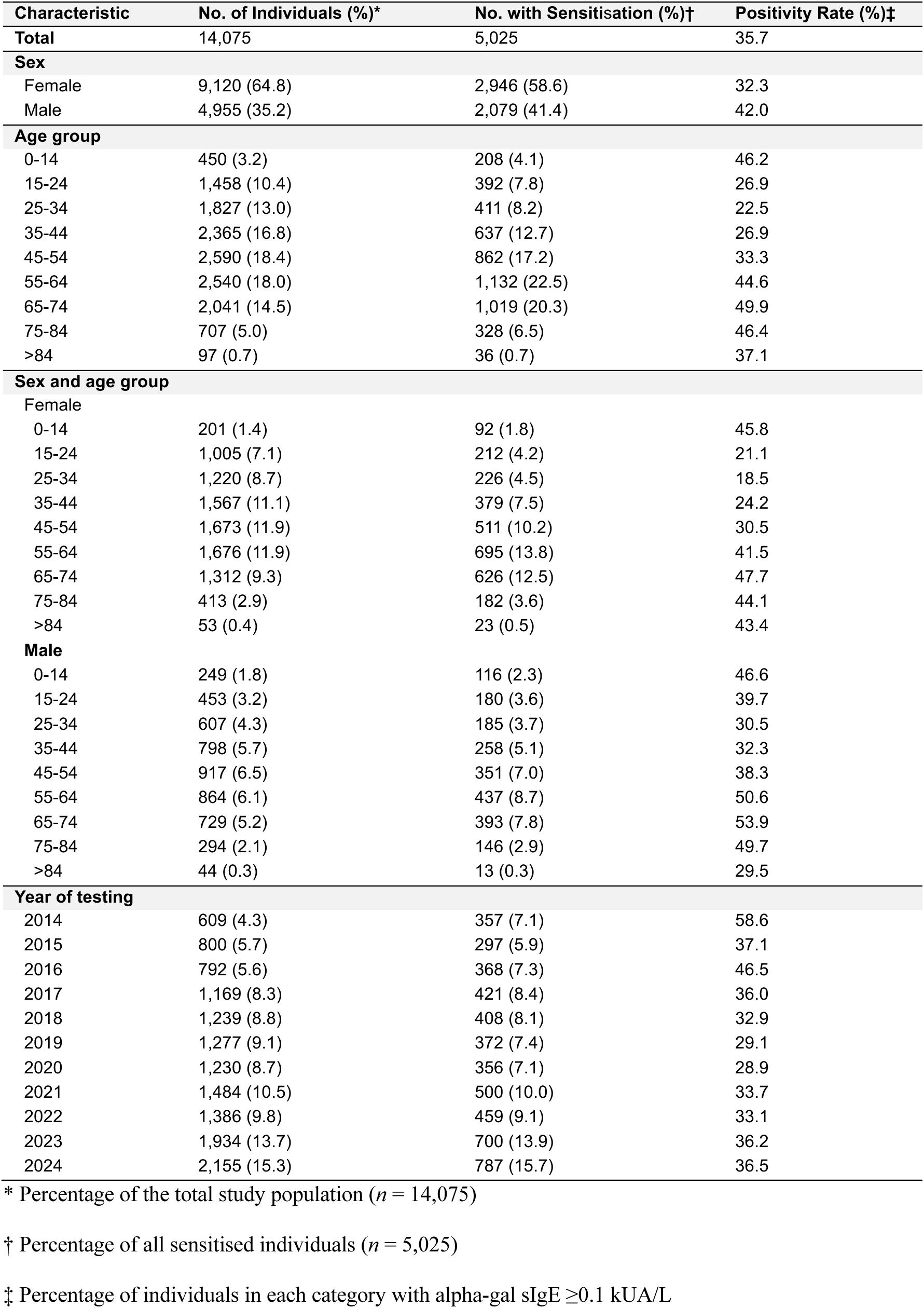
Characteristics of individuals tested for α-Gal sIgE in Australia, January 1, 2014 – December 31, 2024.

**Figure 1.**
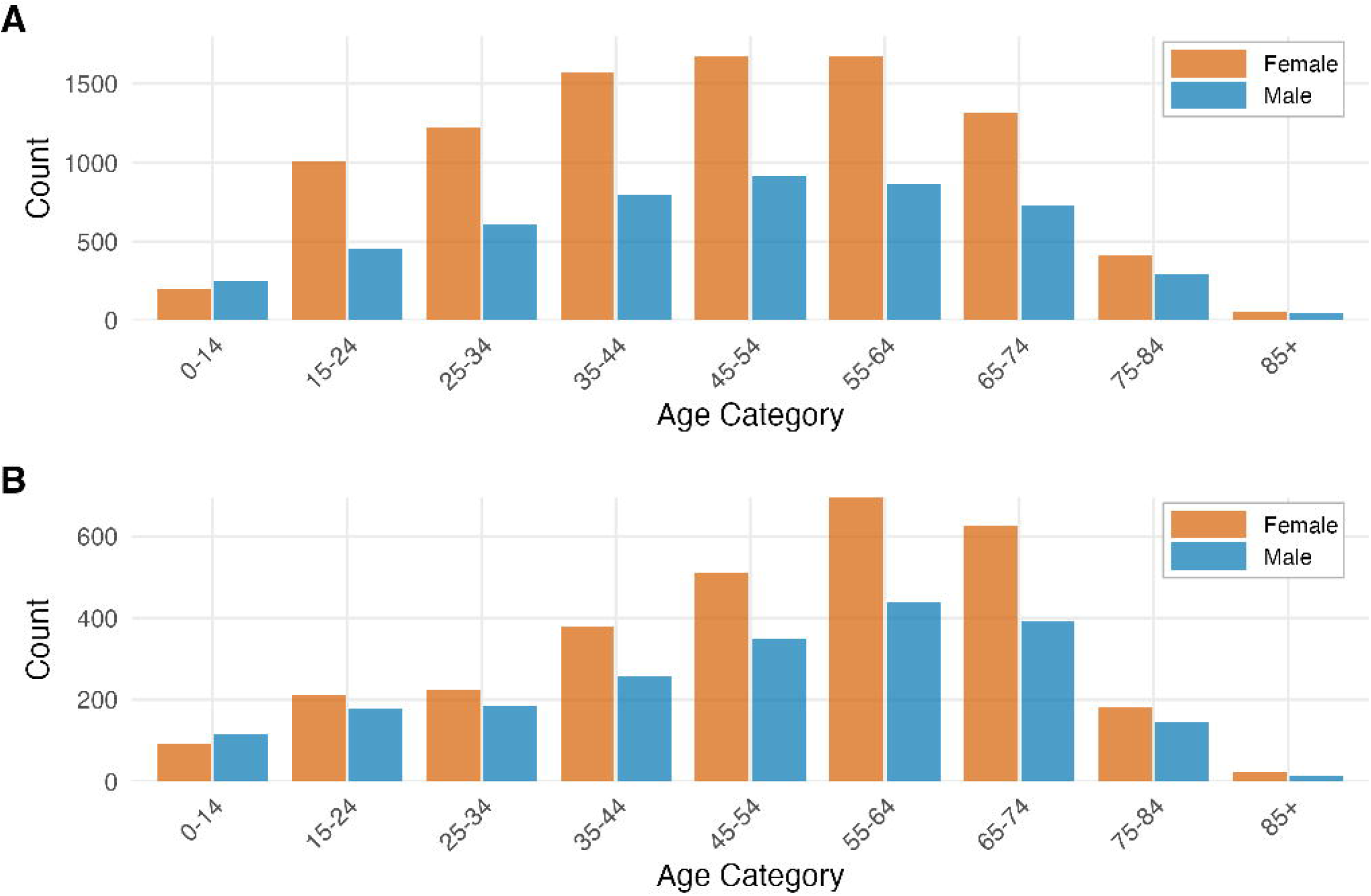
Age distribution by sex of all tested people (A) and people with suspected MMA (B).

Age-and-sex stratified analysis using females aged 25–34 years as reference (18.5% positivity; n = 1,220) revealed males aged 65–74 years as the group with the highest likelihood of returning a positive test (53.9% positivity; RR = 2.91), followed by males aged 55–64 and females aged 65–74 **(Table S1; Figure S1)**. Within each age category, males consistently demonstrated higher positivity rates than females. The 35.4 percentage point difference in positivity between the lowest-positivity group (females 25–34) and the highest-positivity group (males 65–74) suggests that both advancing age and male sex contribute to α-Gal sIgE sensitisation risk among those tested (**Table S1**). However, despite males’ higher positivity rates, females comprised the majority of both tested individuals and positive cases **(Table 1)**. These patterns may reflect both underlying epidemiology and clinical testing behaviour which could result in over-representation of groups with higher perceived clinical suspicion.

### 3.2 Trends in Testing Volume and Case Detection

### Testing volume

Alpha-Gal sIgE testing volume increased 331% from 628 tests in 2014 (609 people) to 2,707 tests in 2024 (2,155 people), with a significant acceleration in 2022 **(Figure 2A**; **Table 1)**. From 2014 to 2021, testing grew steadily at 136.7 tests/year (R² = 0.79; CAGR: 12.5%, CI: 8.5-16.7). However in subsequent years (2022-2024), testing volume grew markedly, increasing by 393 tests/year (R² = 0.82; CAGR: 24.7%, CI: 9.8-41.6). Segmented regression confirmed this 2022 breakpoint as a structural shift in testing practices (SE = 1.26, R² = 0.93, p < 0.01). This growth was driven by two simultaneous processes: geographic expansion (SA3 regions conducting testing increased from 152 to 254) and intensification within existing regions (mean tests per region rose from 4.0 to 8.5 per year) (**Figure 3A-C; Figure S2**). Regions that began testing later (≥2022) had significantly lower positivity rates than early-adopter regions (≤2016; Kruskal–Wallis χ² = 36.8, p < 0.01), suggesting initial testing was concentrated in high-prevalence areas before expanding into lower-burden regions. Detailed decomposition analysis is provided in **Supplementary Information**.

**Figure 2.**
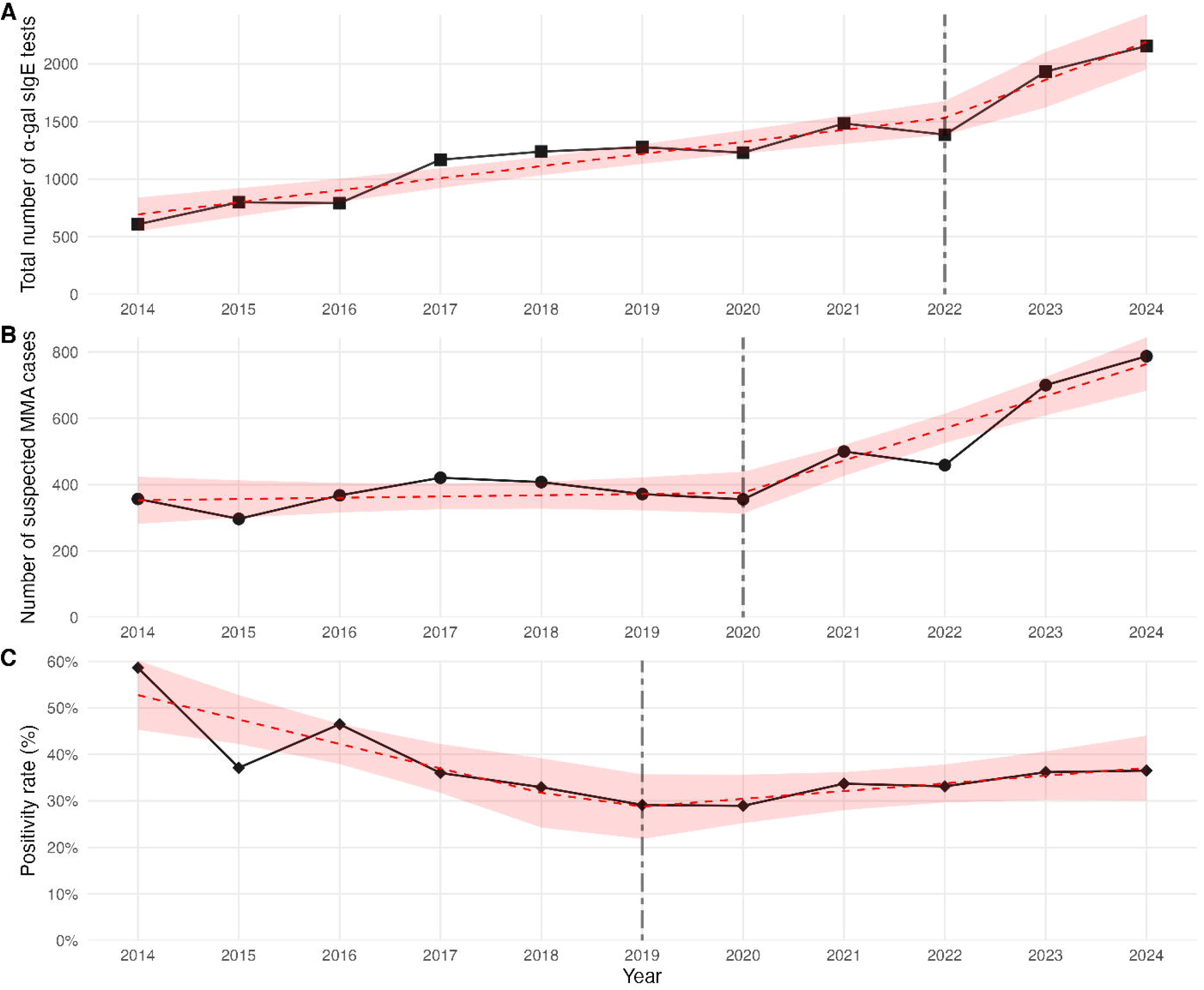
Annual trends in α-Gal sIgE testing in Australia, 2014-2024. (A) Total tests performed. (B) Number of suspected MMA cases. (C) Testing positivity rate. Linear segmented regression models and SE are indicated by red dashed lines and light red areas, and grey dot-dashed line indicated the structural breakpoints identified by the linear segmented regression analysis.

**Figure 3.**
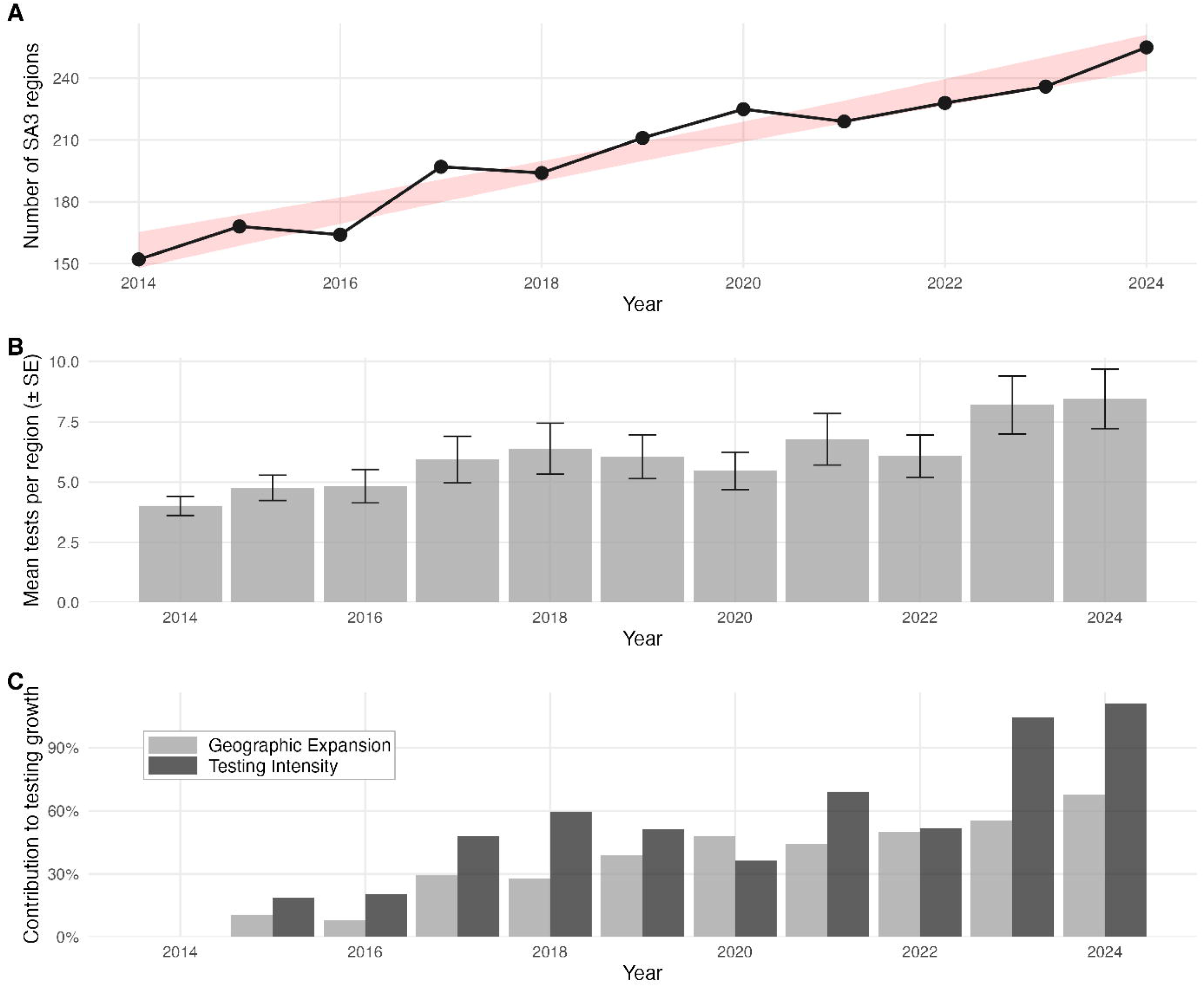
Dual expansion of α-Gal sIgE testing infrastructure in Australia, 2014-2024. (A) Geographic expansion: number of SA3 regions conducting testing; shaded area shows 95% CI. (B) Testing intensity: mean tests per region; error bars show standard error. (C) Relative contributions of geographic expansion and testing intensity to overall testing volume growth.

### Suspected MMA case detections

Suspected MMA case detections increased 120.4% over the study period, from 357 in 2014 to 787 in 2024, with a breakpoint in 2020, two years before the testing volume breakpoint **(Table 1**; **Figure 2B)**. From 2014–2020, annual case numbers remained relatively stable, increasing by just 6.68 cases/year (R² = 0.91). From 2020 onwards, case detection accelerated markedly at 110.2 cases/year (R² = 0.84), representing a 121.1% increase. Testing positivity rates also displayed two phases with a 2019 breakpoint (SE = 1.5, R² = 0.76, p < 0.01; **Figure 2C**): an initial decline from 58.6% (2014) to 29.1% (2019) at −5.2%/year, likely reflecting diagnostic selection bias when testing was largely restricted to patients with high clinical suspicion, followed by an increase to 36.5% in 2024 at 1.2%/year, notably despite a 75.2% rise in testing volume over the same period.

While testing expansion is the dominant driver of case growth, 19–41% of the observed increase could not be attributed to surveillance expansion alone **(Figure S3)**. To quantify this, we decomposed case growth using the 2018–2020 period as the reference baseline, by which time testing had stabilised into a representative referral pattern (mean positivity 30.3%). Negative binomial regression confirmed that a time trend in positivity remained significant after controlling for testing volume (rate ratio = 0.966 per year, CI 0.94–0.99; p < 0.05) (**Supplementary Information**).

### 3.3 Geographic distribution of suspected MMA cases

Suspected MMA cases predominantly occurred along Australia’s east coast within the range of the causative tick *I. holocyclus* **(Figure 4)**. However, cases were recorded from every state and territory, with 166 (3.8%) cases identified outside the tick’s recognised range, likely reflecting exposure during travel or prior residence in endemic areas.

**Figure 4.**
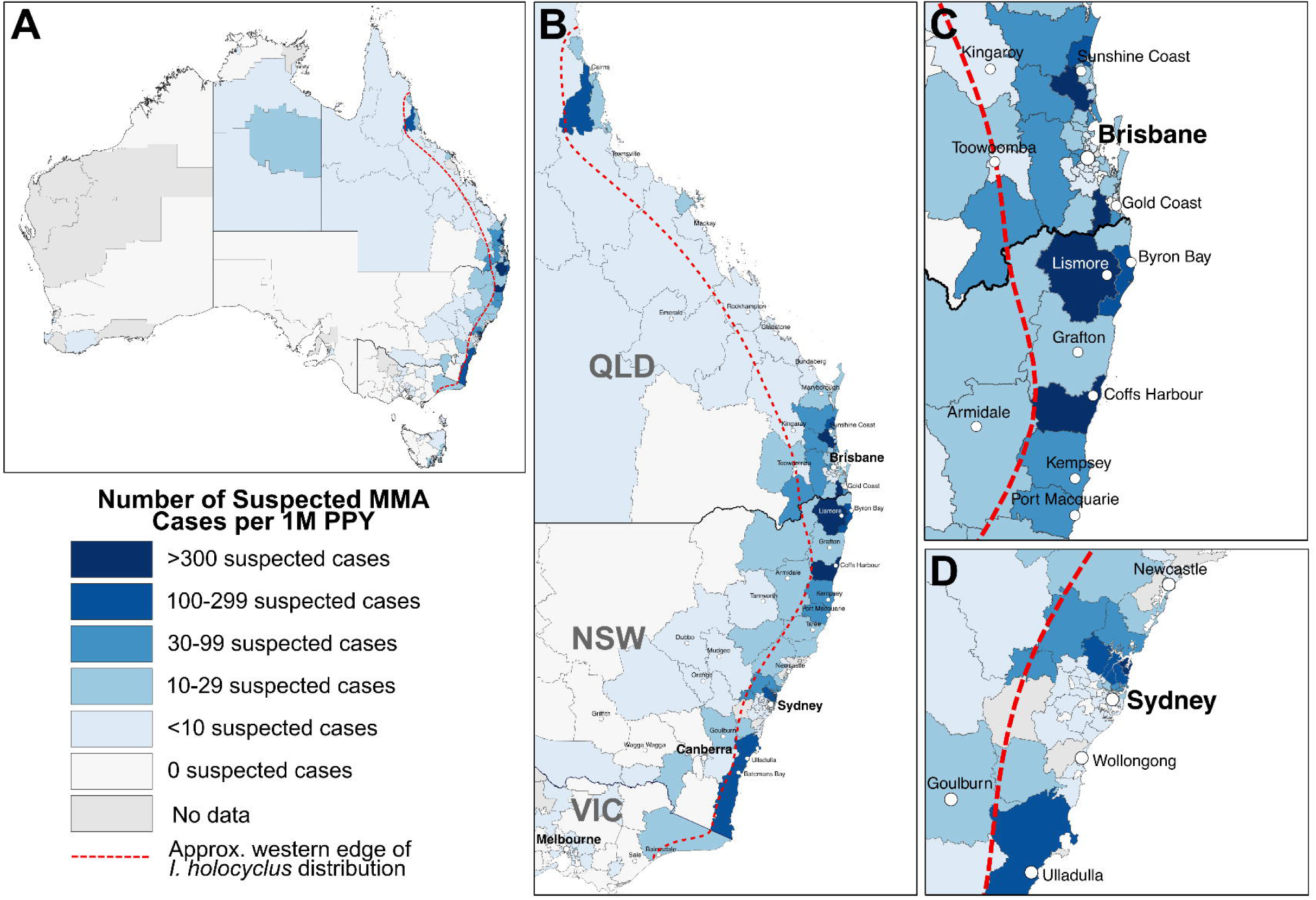
Geographic distribution of suspected mammalian meat allergy cases per million population per year in Australia 2014-2024. (A) all Australia, (B) Eastern Australia, (C) Southeast Queensland and northern New South Wales, (D) Sydney, central coast, and south coast New South Wales. Red dashed line indicated the approximate western edge of the distribution of the causative tick *Ixodes holocyclus*. An interactive version of these maps is available at https://alexandergofton.github.io/Tick-Induced_MMA_in_Australia

Within the *I. holocyclus*-endemic zone, suspected MMA cases showed striking geographic clustering across SA3 regions, with a Gini coefficient of 0.86 (CI: 0.83–0.89). This extreme spatial inequality, approaching the theoretical maximum of 1.0, indicated that the majority of Australia’s MMA burden was concentrated in a small subset of SA3 regions **(Figure 5)**. Spatial autocorrelation analysis confirmed significant positive clustering (Moran’s I: 0.291, p < 0.01), demonstrating that regions with high MMA burdens were geographically proximate rather than randomly distributed. When adjusted for population size, just three SA3 regions accounted for 25.3% of the total MMA burden, 9 regions accounted for 52%, and 20 regions accounted for 70.7% **(Figure 5)**.

**Figure 5.**
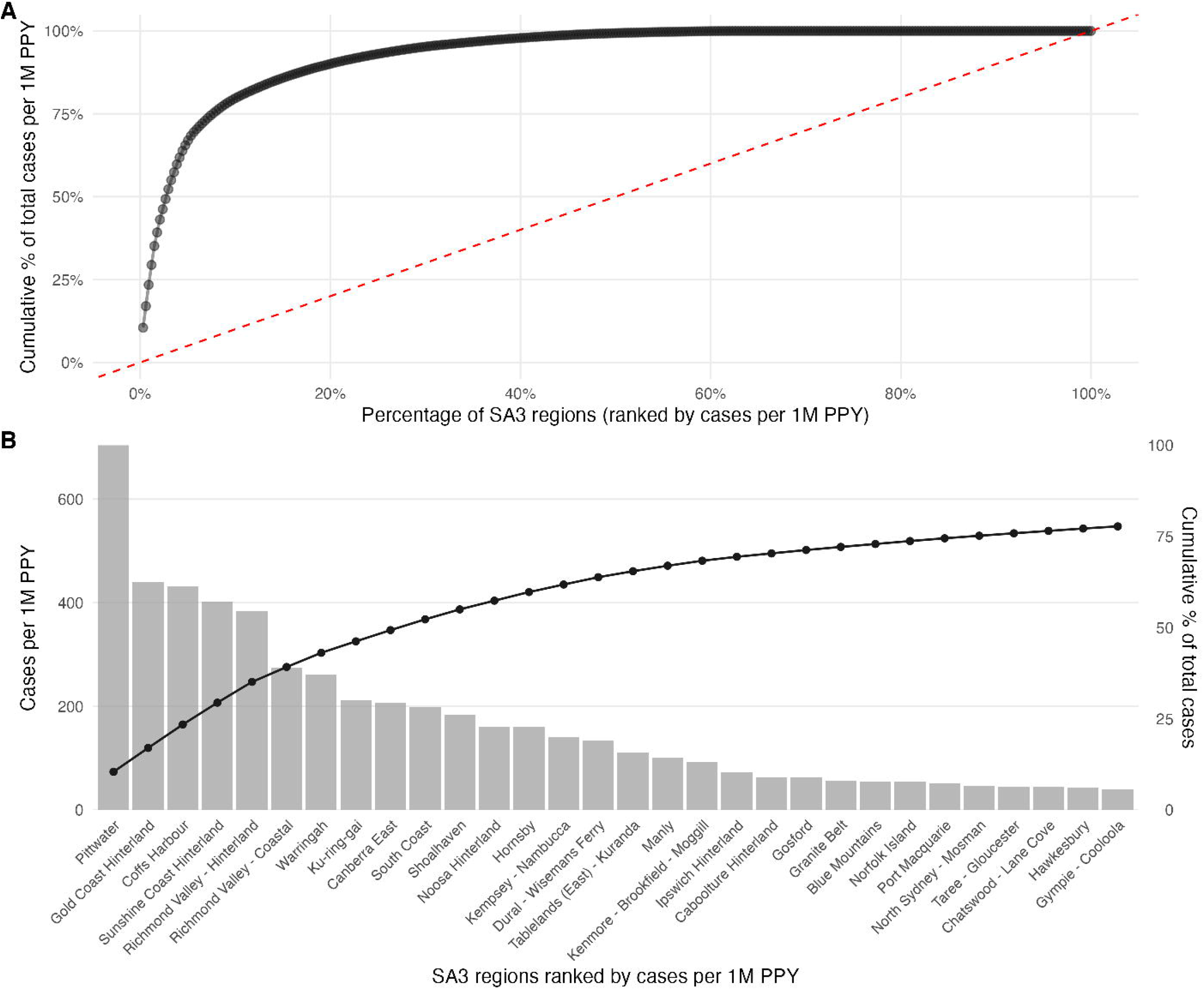
Geographic concentration of suspected alpha-gal syndrome cases across Australian SA3 regions. (A) Lorenz curve showing cumulative distribution of cases across SA3 regions (ranked by case burden). The diagonal dashed line represents perfect equality; deviation from this line indicates concentration. (B) Pareto chart showing the proportion of total cases accounted for by cumulative percentages of SA3 regions.

To assess whether geographic clustering reflected selective referral to high-awareness clinicians rather than true disease distribution, we analysed 6,404 tests with available referring clinician postcodes. Positivity rates were similar between high- and low-volume clinicians, most patients were tested locally (median 14.4 km from residence), and regional variation was not explained by referral patterns, suggesting that case clustering reflects genuine disease burden. Full analysis provided in the **Supplementary Information**.

Pittwater (Sydney’s Northern Beaches, NSW) exhibited the highest MMA burden (684 cases per 1M PPY). Northern Sydney encompassed several contiguous high-burden SA3 regions, including Warringah, Ku-ring-gai, and Hornsby, collectively accounting for 25.7% of all national cases. Other high-burden regions in NSW included Richmond Valley, Coffs Harbour, Gosford, Shoalhaven, and the South Coast. Within Queensland, cases were concentrated in south-east Queensland hinterland regions (Gold Coast, Sunshine Coast), with Kuranda the only high-burden region in the state’s north. Full regional statistics are provided **Supplementary Table S2**.

### 3.4 α-Gal sIgE levels decline over time

Among the 1,515 patients who received multiple α-Gal sIgE tests over the study period, most (67.9%) had just two tests (range: 2–15). The median interval between tests was 1 year (range: 0–10 years), and 1,160 of these patients (76.6%) had at least one positive test result.

Examination of patient-specific α-Gal sIgE trajectories over the course of their testing (fixed effect plus individual random effects) revealed that 93.2% of patients exhibited declining α-Gal sIgE levels over time **(Figure 6A-B)**. Examining cohort-level trends, linear mixed-effects modelling with random intercepts and slopes also demonstrated a significant decline in α-Gal sIgE levels over time (β = −0.142 log kU/L per year, CI: −0.16 to −0.12, p < 0.01), corresponding to an estimated annual reduction of approximately 13.2% of absolute α-Gal sIgE levels **(Figure 6A-B)**. The close agreement between the population-level fixed effect (−0.142) and the median individual slope (−0.124) indicates that most patients followed the average cohort trajectory, with the small difference reflecting individuals with more extreme changes. While the decline in α-Gal sIgE over time was statistically significant, the primary determinant of decline rate was the initial α-Gal sIgE concentration which accounted for 60.9% of the variance in individual decline rates **(Figure 6C)**. Patients with higher baseline in α-Gal sIgE concentrations experienced significantly steeper declines (r = −0.78, CI: −0.80 to −0.76, p < 0.01; **Figure 6C**).

**Figure 6.**
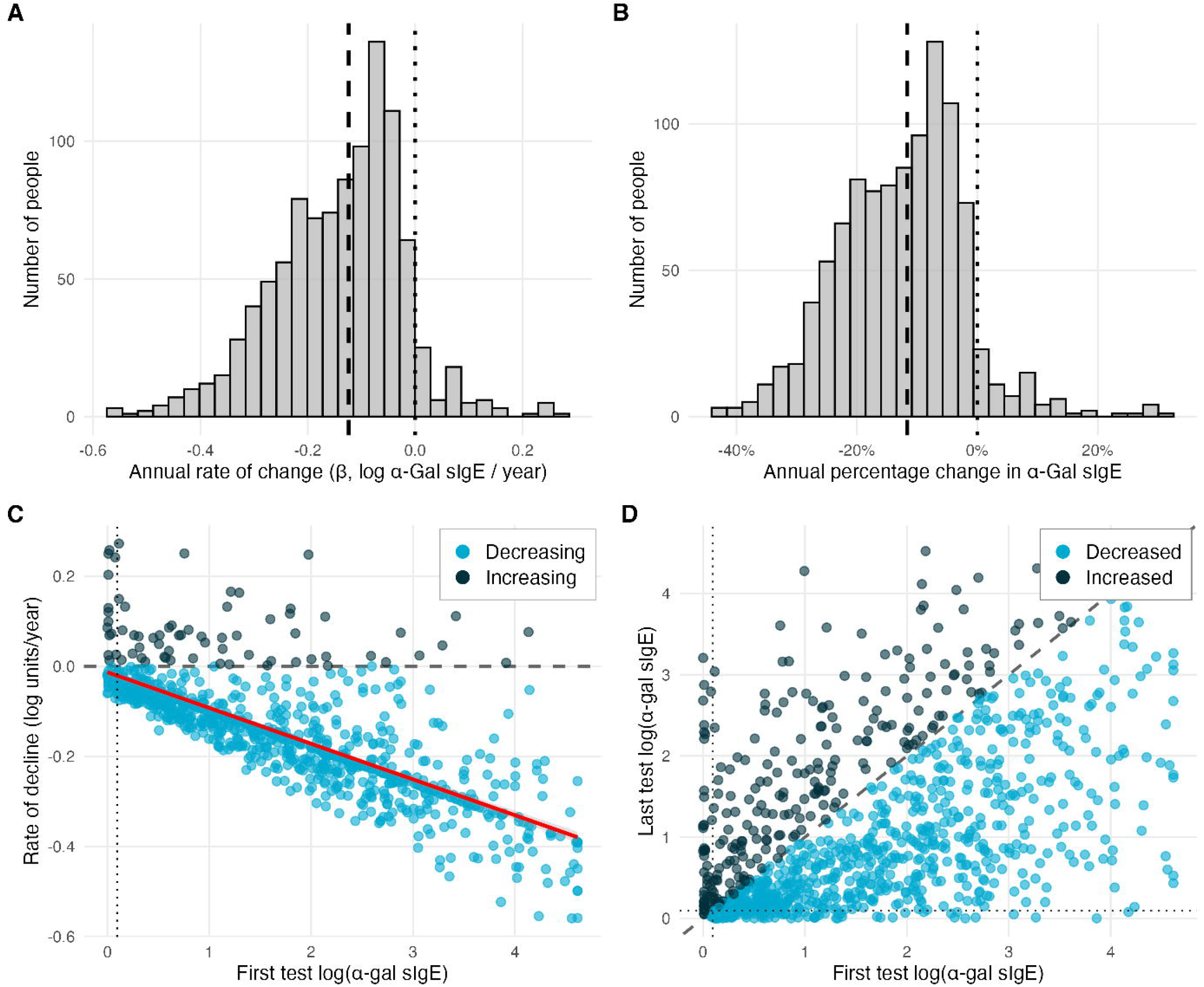
Longitudinal changes in α-Gal sIgE levels in individuals with ≥2 tests. Distribution of individual annual rates of change (β) in log α-Gal sIgE per year in absolute number (A) and as percentage change (B), estimated from linear mixed-effects models. Negative values indicate declining antibody levels. Dotted line indicates no change; dashed line indicates the median rate of change. (C) Relationship between baseline α-Gal sIgE concentration and individual rate of decline. Each point represents one individual. The red line shows the linear regression fit. (D) Comparison of first and last α-Gal sIgE measurements (log-transformed) for each individual. Points below the diagonal dashed line (slope = 1) indicate individuals whose levels decreased between first and last test; points above indicate increases. The red line shows the linear regression fit.

When comparing each patient’s first and final α-Gal sIgE measurements the median decrease in α-Gal sIgE was 2.2 kU/L, representing a 78.8% (CI: 69.2–88.4) reduction from baseline **(Figure 6D)**. Overall, 58.5% of people achieved a ≥50% reduction in their α-Gal sIgE levels within 2.1 years. However, only 9% of patients seroconverted from positive (≥0.1 kU/L) to negative (<0.1 kU/L) during the study period.

## 4. Discussion

This is the first epidemiological assessment of mammalian meat allergy (MMA) in Australia, analysing national pathology data over an 11-year period (2014-2024). Our findings define several core epidemiological features of MMA in the Australian context that are directly relevant to surveillance, clinical recognition, and public health planning. These include a substantial and highly concentrated geographic burden, pronounced demographic risk gradients by age and sex, and rapidly evolving testing and case-detection dynamics.

Both age and sex were strongly associated with MMA risk, suggesting that demographic profiles could inform clinical suspicion and risk-stratification. Females accounted for the majority of individuals tested and were appreciably overrepresented in the total number of suspected MMA cases detected. However, males demonstrated higher positivity rates across most age strata, indicating elevated individual-level risk among males who presented for testing. In parallel, MMA risk increased progressively through adulthood, peaking in individuals aged 45–74 years, a trend which concurs with previous demographic assessments of MMA patients in Australia and internationally [16, 17].

The coexistence of higher testing volumes among females and higher positivity among males underscores the importance of distinguishing population-level case detections from individual-level risk when interpreting this surveillance data. This pattern may reflect sex-specific difference in healthcare-seeking behaviour, with males accessing medical care and diagnostic services less frequently [6, 18, 19]. Therefore, the tested male population may be enriched for more clinically significant cases. The higher seropositivity rate among males and peak risk in middle adulthood is consistent with findings from US and European cohorts [17, 20]. However, though the predominance of females among suspected MMA cases in Australia contrasts with the more balanced sex distribution observed internationally [17, 20, 21], suggesting regional differences in healthcare-seeking behaviour, tick exposure contexts, or public awareness between sexes. Further research examining referral pathways and tick exposure histories could help clarify the factors underlying these demographic patterns.

The geographic distribution of MMA burden in Australia closely mirrors the distribution of the causative tick (*I. holocyclus*) along Australia’s eastern seaboard [9, 10]. Although 3.8% of suspected cases were detected outside of *I. holocyclus*-endemic zones, this is most plausibly explained by travel or prior residence in endemic regions rather than suggesting a broader vector distribution. *Ixodes australiensis* has been reported as a secondary causative species in Australia [22]. However, the paucity of cases in south-west Western Australia (11 suspected cases), where this tick predominates, suggests it induces α-Gal sensitisation less efficiently than *I. holocyclus*, and is a minor contributor to MMA burden in Australia.

Within the *I. holocyclus*-endemic zone, MMA burden demonstrated extreme regional variation, with the vast majority of suspected cases concentrated within a small subset of SA3 regions. Significant spatial autocorrelation further confirmed that high-burden regions cluster geographically, suggesting shared environmental determinants of risk. A parallel pattern is observed in US surveillance data. While suspected cases span a broad, contiguous swath of southern, midwestern, and mid-Atlantic states (consistent with the distribution of the causative tick *Amblyomma americanum*), MMA case density is highly focal, with a small number of counties accounting for a disproportionate share of national burden [17, 20].

*Ixodes holocyclus* thrives in areas with humid microclimates, dense ground cover, and vegetation structure that provides host habitat and protection from desiccation [11, 23, 24]. The concentration of MMA cases in coastal and hinterland regions of NSW and south-east QLD is consistent with these ecological requirements, as these areas provide climatic and vegetative conditions favourable for tick survival [11]. However, *I. holocyclus* habitat suitability alone does not appear to translate directly to MMA burden. In the US, species distribution and risk modelling link MMA risk most strongly to open space development, mixed forest types, and forest edge density, landscape features characteristic of fragmented forest habitats at the peri-urban interface [25, 26]. Similarly, Australia’s highest burden areas likely emerge where favourable ecological conditions coincide with landscape features and patterns of land use that amplify human-tick contact.

Northern Sydney’s high-burden suburbs exemplify this convergence. The area is characterised by remnant and regenerated sclerophyll forest with dense understorey, fragmented by residential development, creating both ideal habitat for tick populations and their native mammalian hosts and frequent opportunities for human-tick encounters in gardens, bushland reserves, and recreational areas [27, 28]. Similar dynamics are evident in Queensland’s hinterland regions, where subtropical rainforest and wet sclerophyll habitats support high tick densities year-round while simultaneously attracting residential development and outdoor recreation at the urban–bushland interface.

The pronounced concentration of MMA burden indicates that geographically targeted interventions may be highly effective in reducing national incidence, with just three SA3 regions accounting for more than one-quarter of Australia’s total burden. Priority strategies may include regional public awareness initiatives, targeted education for healthcare providers, and strategic environmental management in high-risk peri-urban settings. The need for such efforts is underscored by a 2023 Australian survey, which found that only 42% of allergy specialists had access to current educational materials on MMA and just 28% considered their patient communities well-informed [29]. Findings from the 2026 NSW Coronial Inquest into Australia’s first MMA death identified substantial gaps in clinician awareness, diagnostic practices, and anaphylaxis management, and recommended incorporating MMA-specific content into routine emergency department training as well as developing dedicated MMA training modules for frontline clinicians [4].

A critical question in interpreting surveillance data for an emerging condition such as MMA is distinguishing genuine disease emergence from surveillance artefact. Our analysis confirms that testing expansion accounts for the majority (59–81%) of case growth since the 2018–2020 baseline period. However, surveillance artefact alone cannot fully explain the observed trends. The case detection breakpoint preceded the testing volume breakpoint by two years, and positivity rates increased modestly from 2019-2024, despite a 75.2% rise in testing volume, the opposite of the dilution effect expected if case growth were driven purely by broader testing. Taken together, these findings suggest that an increase in real disease burden contributed to the significant rise in suspected MMA detection since 2020, although we cannot definitively rule out alternative factors.

Several climatic and anthropogenic factors known to drive tick-borne disease incidence in other systems may explain these inflection points [25, 30, 31]. Consecutive La Niña years (2020–2023) brought above-average rainfall to eastern Australia, potentially favouring tick survival and increasing abundances [32]. Concurrently, pandemic-related migration from metropolitan centres into regional and peri-urban tick-endemic areas may have increased human-tick encounters [33]. These ecological and demographic shifts occurred alongside growing clinical and public recognition of MMA, which likely contributed independently to increased testing and referral [34]. These factors are not mutually exclusive, and their convergence makes it difficult to isolate any single driver. Ultimately, prospective cohort studies or population-based serosurveys would be required to disentangle surveillance effects from true changes in disease incidence and to clarify the extent to which MMA is undergoing genuine emergence.

Longitudinal analysis of α-Gal sIgE levels in patients with serial testing provides important insights. The finding that 93% of patients showed declining α-Gal sIgE levels over time, with a median annual reduction in α-Gal sIgE of 13.2%, is consistent with observations from US cohorts where tick bite avoidance leads to waning sensitisation [2, 35]. Kim and colleagues reported that α-Gal sIgE in the US decreased in 89% of patients who reported no interval tick bites, with approximately 12% achieving undetectable levels over five or more years of follow-up [35]. Our observation that patients with higher baseline α-Gal sIgE levels experienced steeper absolute declines accords with first-order kinetics of antibody decay [36]. The waning of α-Gal sIgE over time suggests that serial testing may have utility in monitoring disease trajectory. In other IgE-mediated food allergies, evidence increasingly supports the tolerogenic role of continued food ingestion and the efficacy of oral immunotherapy in established allergy [37]. Similar principles may apply to MMA, with emerging evidence suggesting that declining α-Gal sIgE levels could guide safe dietary reintroduction of mammalian foods [38, 39].

### 4.1 Limitations

This study classified positive tests as suspected MMA cases based solely on serological evidence. Not all sensitised individuals develop clinical disease, and we therefore likely overestimate clinical burden while underestimating sensitisation in the untested population. The exact proportion of national testing captured by participating laboratories is unknown, though these represent the major commercial networks conducting α-Gal sIgE testing in Australia at the time of publication. Patient residential postcodes may not reflect tick exposure location, and patients tested at multiple providers would be missed as repeat testers. For longitudinal antibody analysis, patients with serial testing may represent a selected subset with more severe disease or under consistent clinical management, and declines cannot be definitively attributed to tick avoidance. Finally, our findings may not fully represent rural, remote, or underserved populations.

### 4.2 Conclusions

This study provides the first comprehensive epidemiological characterisation of MMA in Australia. Our findings confirm a substantial and geographically concentrated burden, with few SA3 regions accounting for the vast majority of suspected cases nationwide. Age and sex are significant demographic predictors of positivity among those tested, with older people and females disproportionally represented among suspected cases. While testing expansion is the dominant driver of rising case numbers, other factors beyond surveillance expansion are contributing to MMA emergence. These may include increased disease burden due to ecological and demographic changes increasing human-tick contact, and/or improved clinical recognition driving more targeted referral for testing. The observed cohort-level decline in α-Gal sIgE over time further highlights the potential for serial testing to inform clinical monitoring and guide dietary management. Together, these findings provide a foundation for targeted public health responses and underscore the need for prospective studies integrating tick exposure data with individual clinical histories to disentangle surveillance effects from genuine changes in disease incidence.

## End matter

### Artificial Intelligence (AI)

During the preparation of this work the authors used Artificial Intelligence tools under supervision to edit analytical code and to refine scientific writing. All authors reviewed and edited the content and take full responsibility for the content of the published article.

### Data Sharing

The de-identified data analysed here are not publicly available, but requests to the corresponding author for the data will be considered on a case-by-case basis.

