## Supplementary Information for "Tick-Induced Mammalian Meat Allergy in Australia: National Prevalence and Geographic Distribution from Laboratory Surveillance, 2014-2024"

### S1. Supplementary Methods

#### S1.1 Patient demographic and age/sex risk ratios

Age was categorised into 10-year bands: 0-14, 15-24, 25-34, 35-44, 45-54, 55-64, 65-74, 75-84, and 85+ years. These categories were chosen to align with standard demographic reporting and to provide adequate sample sizes within each stratum for risk ratio calculations.

Age distributions between groups were compared using the Wilcoxon rank-sum test (for comparing medians between two groups) and the two-sample Kolmogorov-Smirnov test (for comparing entire distribution shapes). Effect sizes were quantified using Cohen's d, calculated with pooled standard deviation via the *effectsize* package (v1.0.1) [1].

Risk ratios (RR) with 95% confidence intervals were calculated using the *epitools* package (riskratio function) [2]. Risk ratios were computed for: (a) Males versus females (overall and within each age category), (b) Each age group versus the reference group (25-34 years, pooled sexes), (c) All age-sex combinations versus the lowest-risk reference group (females aged 25-34 years). P-values were derived from Fisher's exact test. The Mantel-Haenszel test was used to assess the overall association between sex and positivity after stratifying by age group, with the common odds ratio and 95% CI reported.

#### S1.2 Temporal trends in testing volume and suspected case detection

##### *S1.2.1 Compound annual growth rate*

Compound annual growth rates (CAGR) with 95% confidence intervals were calculated using log-linear regression. For each time series (testing volume, positive cases, positivity rate), a linear model was fitted with log-transformed values as the dependent variable and year as the independent variable. The CAGR was calculated as $(\exp(\beta) - 1) \times 100$, where $\beta$ is the regression coefficient for year. Confidence intervals were derived from the standard error of the year coefficient: $CI = (\exp(\beta \pm 1.96 \times SE) - 1) \times 100$.

##### *S1.2.2 Segmented regression*

Structural breakpoints in temporal trends were identified using segmented linear regression via the *segmented* package in R [3]. This approach fits piecewise linear models and estimates the location(s) of breakpoint(s) where the slope changes significantly. For each outcome (testing volume, case numbers, positivity rate), initial models were specified with one potential breakpoint, and the algorithm iteratively estimated the optimal breakpoint location and 95% confidence interval. Model fit was assessed using $R^2$, and breakpoint significance was evaluated using the Davies test ($p<0.01$ threshold). Separate slopes were estimated for each segment defined by the breakpoints.

*S1.2.3 Decomposition of testing volume growth*

Testing volume growth was decomposed into geographic expansion (new SA3 regions beginning testing) versus testing intensification (increased testing within existing regions). For each year, we calculated: (1) the number of SA3 regions with at least one test, and (2) the mean number of tests per testing region. The contribution of each component was quantified by calculating counterfactual scenarios: volume if only geographic expansion had occurred (with intensity held at baseline), and volume if only intensification had occurred (with geographic coverage held at baseline).

*S1.2.4 Decomposition of growth in suspected MMA cases post 2020*

To quantify the relative contributions of testing volume expansion versus changes in positivity rate to observed case growth, we applied additive decomposition. The total number of cases in any year equals testing volume multiplied by positivity rate. The change in cases from a baseline period can be decomposed into three components:

**Volume effect:** $(\text{Tests}_t - \text{Tests}_0) \times \text{Rate}_0$

**Rate effect:** $\text{Tests}_0 \times (\text{Rate}_t - \text{Rate}_0)$

**Interaction effect:** $(\text{Tests}_t - \text{Tests}_0) \times (\text{Rate}_t - \text{Rate}_0)$

The baseline reference period was chosen as 2018-2020 (pooled), by which time testing had stabilised into a representative referral pattern following the early diagnostic phase characterised by high-suspicion testing (mean positivity 30.3%). Using the 2014 positivity rate (58.6%) as baseline was avoided because this early period likely reflected selection bias toward high-suspicion cases before broader adoption of α-Gal sIgE testing.

To assess whether testing volume alone explained the increase in detected cases, three linear regression models were compared: (1) time only (positive cases ~ year), (2) testing volume only (positive cases ~ total tests), and (3) combined (positive cases ~ year + total tests). Models were compared using $R^2$ and Akaike Information Criterion (AIC). A significant year coefficient in the combined model would indicate a temporal trend in positivity beyond what testing volume alone explains. Additionally, negative binomial regression was fitted to model positive case counts with testing volume as an offset and year as a predictor. The rate ratio for year indicates whether positivity increased (>1) or decreased (<1) over time after accounting for testing volume.

#### S1.3 Geographic distribution of suspected MMA cases

*S1.3.1 Population-adjusted case rates*

Suspected MMA case rates were expressed as cases per million population per year (PPY) for each SA3 region to enable comparison across regions of different population sizes and with published data from other countries. Annual population estimates for each SA3 were obtained from the Australian Bureau of Statistics regional population dataset. The PPY rate was calculated as: PPY = (total incident cases 2014-2024 / sum of annual population 2014-2024) × 1,000,000. This approach accounts for population changes over the study period while providing a stable annualised rate.

*S1.3.2 Gini coefficient for case concentration*

The Gini coefficient was calculated to quantify the inequality of case distribution across SA3 regions. The Gini coefficient ranges from 0 (perfect equality: cases distributed proportionally across all regions) to 1 (perfect inequality: all cases concentrated in a single region). Bootstrap resampling (1000 iterations) was used to estimate 95% confidence intervals.

*S1.3.3 Spatial autocorrelation*

Global spatial autocorrelation was assessed using Moran's I statistic. Spatial weights were constructed using queen contiguity (regions sharing any boundary). P-values were derived from permutation testing (999 permutations).

*S1.3.4 Clinician referral pattern analysis*

To assess whether geographic clustering reflected differential diagnostic recognition by high-volume clinicians rather than true disease distribution, we analysed the subset of tests with available referring clinician postcodes (n=6,404). Clinician postcodes were mapped to SA2 regions, which provide finer geographic resolution than SA3 and are more appropriate for characterising local referral patterns. We first examined whether positivity rates varied systematically by clinician testing volume, which would suggest that high-volume clinicians might have developed greater diagnostic acumen or more selective referral criteria. Clinicians were stratified into volume quintiles based on total referrals, and positivity rates across strata were compared using the Kruskal-Wallis test. Second, we characterised patient travel distances by calculating geodesic distances between patient residential SA2 centroids and referring clinician SA2 centroids using the *sf* package [4]. If patients were travelling substantial distances to consult specific high-awareness clinicians, this could artificially concentrate cases in clinician-dense urban areas rather than reflecting true residential disease burden. We also identified potential referral hubs by calculating the proportion of each clinician SA2's cases that originated from patients residing in different SA2 regions, with hubs defined as SA2 regions where the majority of referred patients resided elsewhere.

*S1.3.5 Mapping and visualisation*

Maps were generated using the *sf* and *ggplot2* packages in R [4, 5]. SA3 shapefiles were obtained from the ABS ASGS Edition 3 geographic boundaries. Geometries were simplified using st_simplify() with dTolerance=100 to reduce file size while preserving boundary accuracy. PPY rates were categorised into bins based on Jenks natural breaks: 0, <10, 10-29, 30-99, 100-299, and >300 suspected cases per million PPY.

#### S1.4 Longitudinal Antibody Dynamics

*S1.4.1 Cohort definition*

Longitudinal analyses included all patients with two or more α-Gal sIgE tests during the study period (n=1,515). Tests were ordered chronologically within each patient and assigned sequential test numbers. The median number of tests per patient was 2 (range 2-17), with 67.9% having exactly two tests and the remainder having 3-17 tests. Among this cohort, 1,160 patients (76.6%) had at least one positive test result.

*S1.4.2 Mixed-effects modelling*

α-Gal sIgE values were log-transformed [log(kU/L + 1)] to normalise distributions and stabilise variance. Linear mixed-effects models were fitted using the lme4 package [6] with the following structure:

$$\log(\alpha\text{-Gal sIgE}) \sim \text{years since first test} + (\text{years since first test} \mid \text{patient ID})$$

This model includes a fixed effect for time (years since first test), representing the average rate of change across the cohort, random intercepts for each patient, allowing different baseline α-Gal sIgE level, and random slopes for each patient, allowing individual variation in the rate of change over time. The fixed effect coefficient (β) represents the average annual change in log(α-Gal sIgE). The percentage annual change was calculated as: $(1 - \exp(\beta)) \times 100$.

Patient-specific rates of change were extracted from the model as the sum of the fixed effect plus each patient's random slope. The distribution of individual slopes was examined to characterise heterogeneity in α-Gal sIgE dynamics. Patients were classified as having declining, stable, or increasing levels based on their individual slopes.

*S1.4.3 First versus last test comparison*

For patients with at least two tests spanning more than zero years, paired comparisons were made between first and last α-Gal sIgE measurements. Paired t-tests were performed on both raw and log-transformed values. The Wilcoxon signed-rank test was used as a non-parametric alternative. Effect size for the Wilcoxon test was calculated as $r = |Z|/\sqrt{N}$, where Z was approximated from the p-value and N is the sample size. Percentage change from baseline was calculated for each patient: ((last - first) / first) × 100.

*S1.4.4 Predictors of decline rate*

Correlation between baseline (first test) α-Gal sIgE concentration and rate of decline was assessed using Pearson correlation on log-transformed values. Linear regression was used to quantify the proportion of variance in decline rates explained by baseline concentration ($R^2$).

*S1.4.5 Seroreversion analysis*

Among patients who had at least one positive test (≥0.1 kU/L), we calculated the proportion who seroreverted (final test <0.1 kU/L). Time to seroreversion was not formally modelled due to the variable follow-up intervals and right-censoring; instead, descriptive statistics on the proportion achieving seroreversion and the median time to 50% reduction were reported.

*S1.4.6 Time to significant decrease*

The time to achieve a clinically meaningful decrease in α-Gal sIgE was analysed by identifying the first test at which each patient achieved ≥50% reduction from their baseline value. The proportion of patients achieving this threshold and the median time to achievement were calculated. A binomial test was used to assess whether the proportion of patients with declining versus increasing α-Gal sIgE levels differed significantly from 50%.

### S2. Supplementary Results

##### S2.1 Testing volume decomposition analysis

The substantial increase in testing volume over the study period was associated with two distinct but simultaneous processes: geographic expansion and intensification of testing **(Figure 3A-C)**. The number of SA3 regions conducting testing expanded from 152 (2014) to 254 (2024), representing a 67.8% increase in geographic reach (Spearman ρ: 0.75, CI: 0.28–0.93, $p < 0.01$; **Figure 3**). Regions that began testing later (≥2022) had significantly lower positivity rates compared to early adopter regions (≤2016; Kruskal–Wallis $\chi^2 = 36.8$, $p < 0.01$), suggesting that initial testing was concentrated in high-prevalence areas before expanding into lower-burden regions.

Concurrently, testing intensity within regions increased substantially over the study period, with the mean number of tests per SA3 region per year rising from 4.0 in 2014 to 8.5 in 2024 (110.9% increase; Spearman $\rho = 0.90$, CI: 0.69–0.97, $p < 0.01$; **Figure 3**). However, this intensification was highly heterogeneous across regions, with a median increase of 66.7% (IQR = 200%; CV = 192%; Levene's test $p < 0.01$; **Figure S2**).

Decomposition analysis using 2014 as the baseline revealed that geographic expansion alone would have increased national testing volume by 67.8%, while intensification within the original 152 regions alone would have increased it by 111%. The observed increase (120.4%) exceeded either component in isolation, with the interaction between geographic expansion and regional intensification accounting for 89.1% of total growth in testing volume **(Figure S2)**. These findings indicate that increasing testing has been driven by both expanding the geographic footprint of testing and increasing test utilisation within established MMA-endemic regions. Geographic expansion likely reflects previously unrecognised disease burden in regions not previously associated with MMA, while intensification within long-standing MMA-endemic regions may reflect a combination of heightened clinical awareness, expanded testing indications, and genuine increases in disease occurrence.

##### S2.2 Suspected MMA case detection decomposition analysis

To disentangle the contributions of testing expansion increases in suspected MMA case detections, we decomposed case growth using the 2018–2020 period as the reference baseline, by which time testing had stabilised into a representative referral pattern (mean positivity 30.3%). Relative to this baseline, the 2024 case count increased by approximately 408 cases, of which 59–81% is attributable to increased testing volume, with the range reflects uncertainty in attribution of the interaction between volume and positivity components **(Figure S3)**. Negative binomial regression confirmed that a time trend in positivity remained significant after controlling for testing volume (rate ratio = 0.966 per year, CI 0.94–0.99; $p < 0.05$), and a log likelihood test confirmed that testing volume alone could not explain increased in case detections (p<005). These results indicate that while increased testing volume is the dominant driver of case growth, approximately 19–41% of the observed case increase between the 2018–2020 baseline period and 2024 could not be attributed to surveillance expansion **(Figure S3)**.

##### S2.3 Clinician postcode analysis

To assess whether geographic clustering of suspected MMA cases was influenced by selective referral to high-awareness clinicians rather than true disease distribution, we analysed 6,404 tests with available referring clinician postcodes. High-volume clinicians (≥50 tests total) showed similar positivity rates to lower-volume clinicians (39.7% vs 36.5%; Mann–Whitney $p > 0.01$), and clinician testing volume was not correlated with positivity rate (Spearman r = 0.15, $p > 0.01$). Most patients (77%) underwent testing within 50 km of their residence (median 14.4 km), indicating predominantly local access to diagnostic services. Together, these patterns suggest that regional variation in case numbers reflects genuine disease burden rather than clinician awareness or referral-driven diagnostic bias. Several high-volume diagnostic hubs were located outside regions of high case density, however, these were revealed to be major healthcare facilities (eg. major hospitals) with established tertiary referral services, rather than centres attracting disproportionate MMA case referrals.

### Supplementary Tables and Figures

**Table S1** Individual demographic risk factors among those tested for α-Gal sIgE in Australia, January 1, 2014 – December 31, 2024.

| Characteristic | No. Sensitized / No. Tested | Risk Ratio (95% CI) | Significance* |
|---|---|---|---|
| **Sex** | | | |
| **Female†** | 2,946 / 9,120 | Reference | — |
| **Male** | 2,079 / 4,955 | 1.30 (1.24-1.36) | * |
| **Age group** | | | |
| 0-14 | 208 / 450 | 1.02 (0.85-1.21) | NS |
| 15-24 | 392 / 1,458 | 1.31 (1.21-1.42) | * |
| 25-34† | 411 / 1,827 | 1.17 (1.10-1.24) | * |
| 35-44 | 637 / 2,365 | 1.12 (1.06-1.18) | * |
| 45-54 | 862 / 2,590 | 1.13 (1.06-1.19) | * |
| 55-64 | 1,132 / 2,540 | 1.18 (1.09-1.28) | * |
| 65-74 | 1,019 / 2,041 | 1.13 (1.03-1.25) | * |
| 75-84 | 328 / 707 | 1.11 (0.96-1.28) | NS |
| >84 | 36 / 97 | 0.80 (0.59-1.09) | NS |
| **Sex and age group** | | | |
| **Female** | | | |
| 0-14 | 92 / 201 | 2.47 (2.04-2.99) | * |
| 15-24 | 212 / 1,005 | 1.14 (0.96-1.35) | NS |
| 25-34† | 226 / 1,220 | Reference | — |
| 35-44 | 379 / 1,567 | 1.31 (1.13-1.51) | * |
| 45-54 | 511 / 1,673 | 1.65 (1.44-1.89) | * |
| 55-64 | 695 / 1,676 | 2.24 (1.96-2.55) | * |
| 65-74 | 626 / 1,312 | 2.58 (2.26-2.94) | * |
| 75-84 | 182 / 413 | 2.38 (2.03-2.79) | * |
| >84 | 23 / 53 | 2.34 (1.69-3.26) | * |
| **Male** | | | |
| 0-14 | 116 / 249 | 2.51 (2.11-3.00) | * |
| 15-24 | 180 / 453 | 2.14 (1.82-2.53) | * |
| 25-34 | 185 / 607 | 1.65 (1.39-1.95) | * |
| 35-44 | 258 / 798 | 1.75 (1.50-2.04) | * |
| 45-54 | 351 / 917 | 2.07 (1.79-2.39) | * |
| 55-64 | 437 / 864 | 2.73 (2.39-3.12) | * |
| 65-74 | 393 / 729 | 2.91 (2.54-3.33) | * |
| 75-84 | 146 / 294 | 2.68 (2.27-3.16) | * |
| >84 | 13 / 44 | 1.59 (0.99-2.55) | NS |

†Reference group; * = $p < 0.01$, NS = not significant

**Table S2** Regional distribution of suspected MMA cases in Australia, 2014–2024.

| SA3 region | Number of suspected MMA cases (2014-2024) | Suspected cases per 1M PPY |
|---|---|---|
| Pittwater | 479 | 684.0 |
| Gold Coast Hinterland | 96 | 440.0 |
| Sunshine Coast Hinterland | 251 | 402.8 |
| Richmond Valley - Hinterland | 301 | 381.6 |
| Coffs Harbour | 348 | 348.3 |
| Richmond Valley - Coastal | 258 | 275.3 |
| Warringah | 408 | 234.0 |
| Ku-ring-gai | 266 | 192.9 |
| South Coast | 154 | 187.7 |
| Noosa Hinterland | 42 | 160.3 |
| Hornsby | 140 | 147.3 |
| Shoalhaven | 166 | 143.2 |
| Dural - Wisemans Ferry | 34 | 111.6 |
| Tablelands (East) - Kuranda | 52 | 110.8 |
| Kenmore - Brookfield - Moggill | 49 | 92.4 |
| Manly | 40 | 81.8 |
| Kempsey - Nambucca | 44 | 79.1 |
| Ipswich Hinterland | 54 | 73.2 |
| Caboolture Hinterland | 10 | 62.3 |
| Granite Belt | 26 | 56.8 |
| Gosford | 111 | 56.5 |
| Blue Mountains | 47 | 54.4 |
| North Sydney - Mosman | 49 | 45.2 |
| Hawkesbury | 12 | 43.3 |
| Chatswood - Lane Cove | 54 | 41.3 |
| Gympie - Cooloola | 23 | 39.7 |
| The Gap - Enoggera | 22 | 37.0 |
| Port Macquarie | 33 | 35.6 |
| Bribie – Beachmere | 14 | 35.2 |
| Mudgeeraba - Tallebudgera | 12 | 30.5 |
| Lake Macquarie – West | 27 | 29.9 |

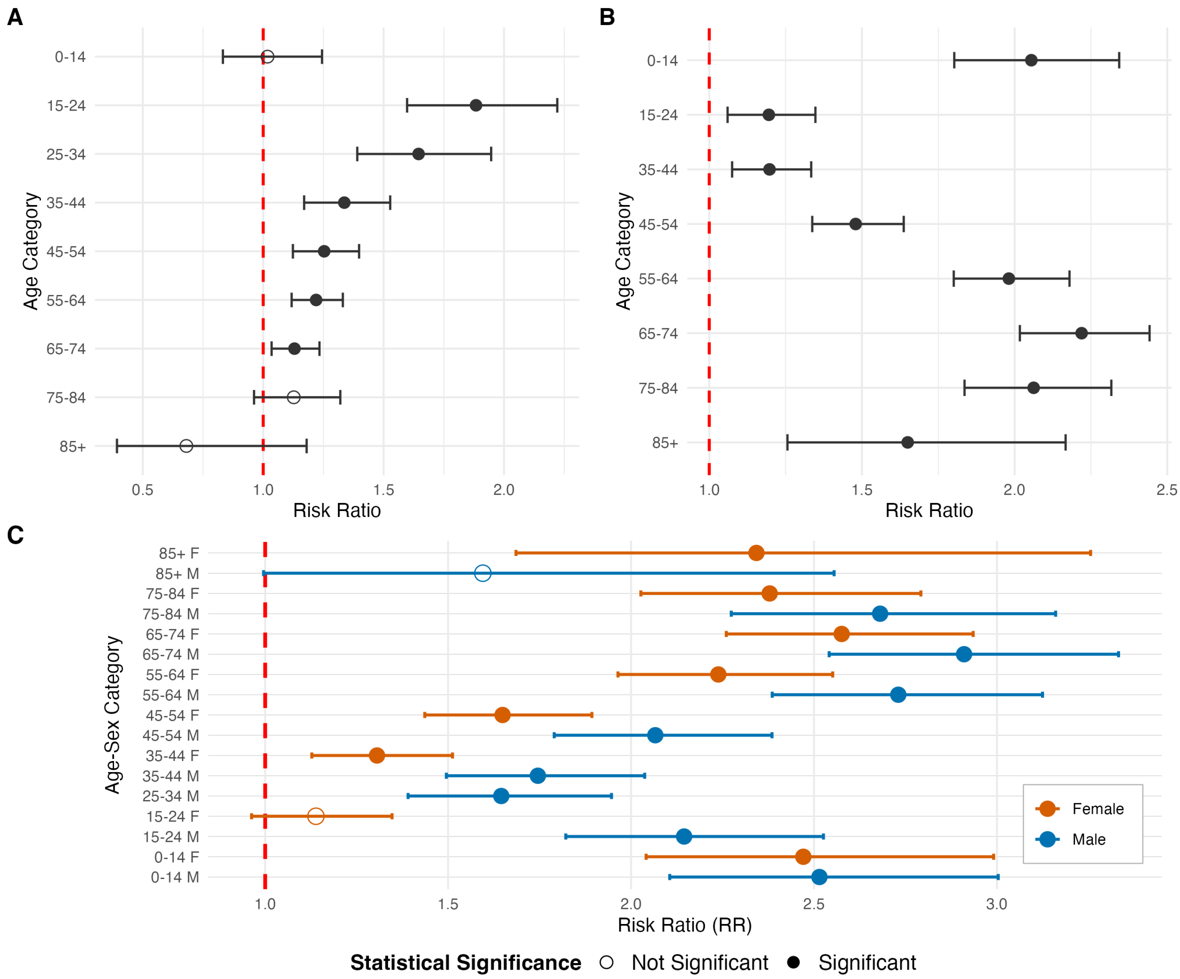


**Figure S1** Risk ratios comparing: (A) males to females within each age category, (B) each age group to the reference (25–34 years, pooled sexes), and (C) all age-sex combinations to females aged 25–34 years (the lowest-risk group, 18.5% positivity). Filled circles indicate statistical significance ($p < 0.01$); open circles indicate non-significant comparisons. Error bars represent 95% confidence intervals. Dashed vertical line indicates RR = 1.0 (no difference from reference group).

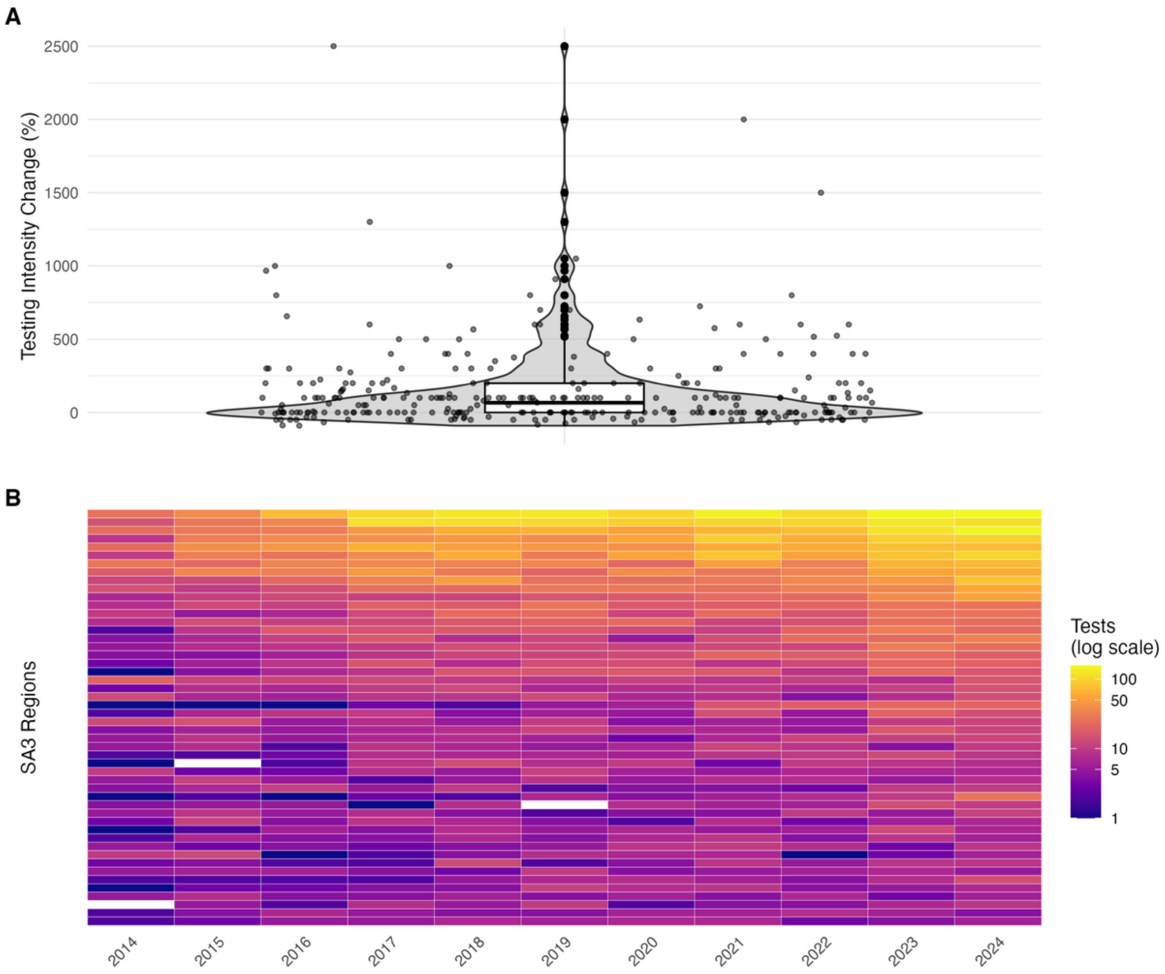


**Figure S2** Dual expansion of α-Gal sIgE testing infrastructure in Australia, 2014-2024. (A) Geographic expansion: number of SA3 regions conducting testing; shaded area shows 95% CI. (B) Testing intensity: mean tests per region; error bars show standard error. (C) Relative contributions of geographic expansion and testing intensity to overall testing volume growth (D) Heterogeneity in regional testing intensity changes over the study period; violin plot shows probability density, boxplot indicates median (66.7%) and interquartile range (0-200%), and individual points represent SA3 regions. (E) Heatmap showing annual testing volume for the 50 SA3 regions with highest total test numbers from 2014-2024 in descending order.

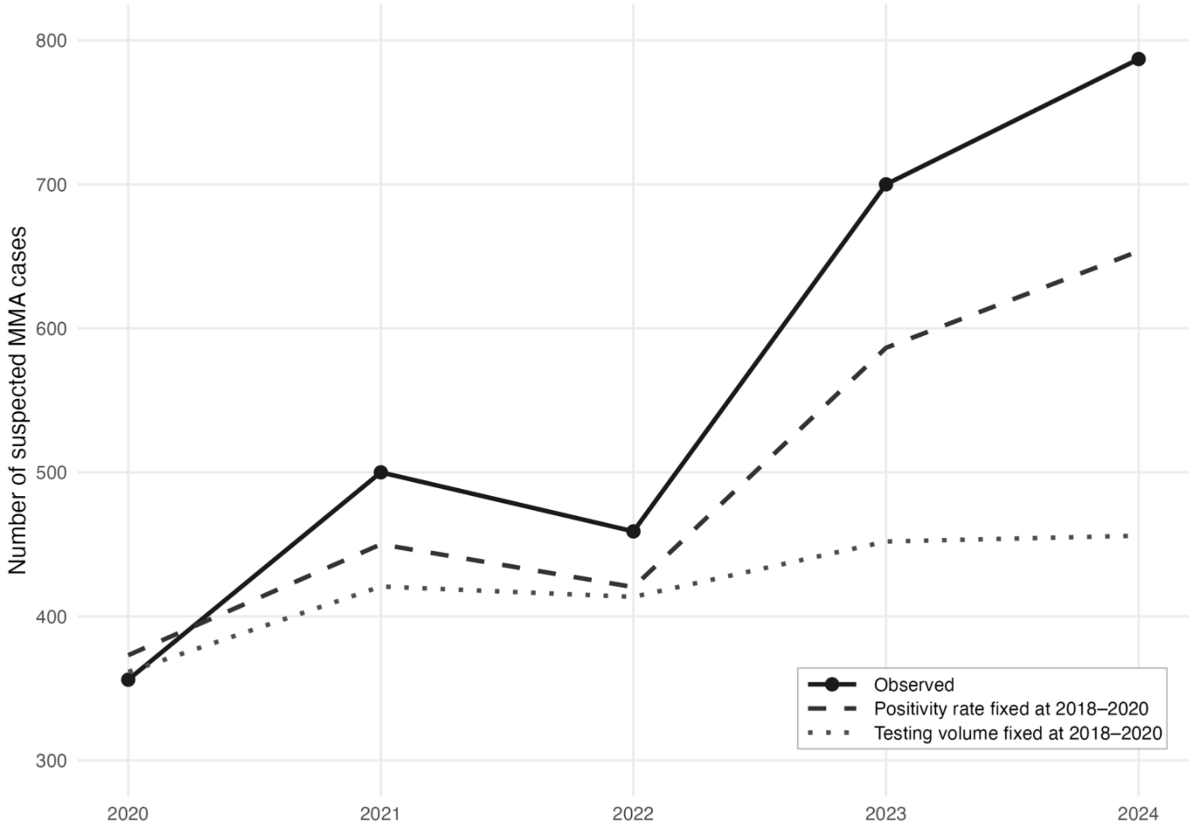


**Figure S3** Counterfactual decomposition of suspected MMA case counts, 2020–2024. Observed annual case counts (solid line with points) compared against two counterfactual scenarios to decompose the drivers of post-2020 case growth. The 2018–2020 baseline represents the pre-expansion period characterised by relatively stable testing volume and positivity rates.
